# Machine Learning-based Mortality Prediction for Pediatric Fulminant Myocarditis Using Cytokine Profiles

**DOI:** 10.1101/2025.05.07.25327044

**Authors:** Sihuan Jing, Takanori Suzuki, Yoji Nomura, Katsuyuki Kunida, Yuichi Sakumura, Hidetoshi Uchida, Kazuyoshi Saito, Ryoichi Ito, Machiko Kito, Satoru Kawai, Kenta T. Suzuki, Alejandro A. Floh, Junichiro Yoshimoto, Tetsushi Yoshikawa, Kazushi Yasuda

**Author notes:** **Corresponding Author**: Yoji Nomura, MD, Katsuyuki Kunida, PhD, Yuichi Sakumura, PhD, Department of Cardiology, Aichi Children’s Health and Medical Center, 426-7, Morioka-cho, Obu, Aichi :474-8710, Japan, facsimile; Department of Computational Biology, Fujita Health University School of Medicine, 1-98 Dengakugakubo Kutsukake cho, Toyoake, Aichi 470-1192, Japan, facsimile; Graduate School of Science and Technology/ NAIST Data Science Center, Nara Institute of Science and Technology, 8916-5 Takayama, Ikoma, NARA 630-0192 JAPAN. **Contributed equally to this work**: Sihuan Jing and Takanori Suzuki.

## Abstract

**Background:** Fulminant myocarditis (FM) is a rare but life-threatening pediatric condition that rapidly progresses to cardiogenic shock and fatal arrhythmias. Early identification of prognostic biomarkers is vital for timely intervention and better outcomes. Although inflammatory cytokines contribute to FM pathogenesis, their prognostic value remains unclear. This study aimed to identify mortality-associated markers by integrating cytokine profiles and clinical variables through a machine learning approach.

**Methods:** We retrospectively analyzed 21 pediatric FM cases from two tertiary centers (2012–2022). At admission, 37 cytokines and 14 clinical parameters were assessed. Partial least squares discriminant analysis was employed to identify prognostic features, with variable importance in projection scores quantifying their contribution. Model performance was evaluated using leave-one-out cross-validation. Statistical significance was determined via the Benjamini-Hochberg method at a false discovery rate of 0.05.

**Results:** Of the 51 features analyzed, 23 emerged as key predictors with variable importance in projection scores above 1.0, including 20 cytokines and three clinical parameters. Six cytokines (TNF-α, M-CSF, MIP-1α, IL-8, IL-6, and IL-15) were both statistically significant and highly important. Elevated CK-MB and lactate levels and lower pH were also linked to poor outcomes. The model performed robustly, with an AUC of 0.92, 85.7% accuracy, 92.9% sensitivity, and 71.4% specificity.

**Conclusions:** TNF-α emerged as a key cytokine linked to mortality in pediatric FM, supporting its role as a prognostic biomarker and potential therapeutic target.

## Introduction

Fulminant myocarditis (FM) is characterized by extensive and severe myocardial inflammation, frequently resulting in fatal outcomes due to malignant arrhythmias and cardiogenic shock. Although advances in mechanical circulatory support (MCS) and cardiac transplantation have improved care, FM remains associated with high mortality risk, largely due to diagnostic delays and suboptimal timing of intervention, highlighting the importance of early recognition and treatment **[1, 2]**. A prior study investigating prognostic indicators of severe outcomes in pediatric FM identified elevated B-type natriuretic peptide (BNP) levels, reduced left ventricular ejection fraction (LVEF), and acidosis as predictors of poor prognosis **[3, 4]**. However, this study had some limitations, including its single-center, retrospective nature, inconsistent definitions of FM, absence of myocardial biopsy-confirmed cases, and reliance on univariate analysis alone.

Cytokines play a critical role in the pathophysiology of myocarditis, with tumor necrosis factor-alpha (TNF-α) and IL-6 recognized as established markers of disease severity **[5, 6]**. In our previous study, we used machine learning to show that multiple cytokines can stratify the severity of acute myocarditis (AM) and FM in children at the time of admission **[7]**. However, the prognostic utility of cytokine profiles in predicting outcomes in fatal FM remains unclear. This study aims to identify predictors of poor prognosis in pediatric FM, a rare but highly severe condition through the integration of clinical characteristics and cytokine profiling.

Medical applications of machine learning have advanced the analysis of high-dimensional clinical datasets **[8, 9]**. These methods can uncover latent associations, enable robust classification, and improve predictive precision in clinical research **[10]**. Previously, partial least squares discriminant analysis (PLS-DA) and variable importance in projection (VIP) scores were employed for feature selection, which enhanced the identification of cytokine markers linked to patient outcomes in pediatric asthma **[8, 11]**. By integrating cytokine profiling with machine learning, this study seeks to improve diagnostic accuracy, elucidate the inflammatory landscape of FM, and identify key biomarkers for potential therapeutic targeting.

## Methods

### Patients

Patients with FM admitted to Aichi Children’s Health and Medical Center and Fujita Health University between January 2012 and December 2022 were retrospectively analyzed. AM was defined by at least one of the following criteria within 30 days of symptom onset: (1) histopathological evidence of inflammatory cell infiltration and myocardial injury on biopsy or (2) clinical presentation consistent with heart failure, characterized by a progressive course and elevated serum high-sensitivity troponin or creatine kinase MB (CK-MB) levels **[2]**. FM was characterized by low cardiac output syndrome requiring MCS due to cardiogenic shock, life-threatening ventricular tachycardia (VT), or bradyarrhythmia such as complete atrioventricular block (CAVB) **[1, 12]**. Of the 21 patients included, 12 (57.1%) were diagnosed via myocardial biopsy, while the remaining cases were diagnosed clinically. This study received approval from the institutional review boards of Aichi Children’s Health and Medical Center and Fujita Health University (Approval No. 2019027 and HM21-575). Informed consent was obtained through an opt-out process in accordance with institutional guidelines, ensuring appropriate disclosure to all patients or guardians.

### Clinical data

The clinical data assessed in this study included sex, age, height, body weight, body surface area, arterial blood gas pH, lactate, BNP, troponin T, CK-MB, and LVEF. The interval from symptom onset to hospital admission (days) was also documented, along with the presence of VT and CAVB.

### Cytokine profile

A total of 37 cytokines were measured using the same methodology described in our previous study. The panel included analytes such as EGF, eotaxin, G-CSF, GM-CSF, IFN-α, IFN-γ, IL-1α, IL-1β, IL-1RA, IL-2, IL-3, IL-4, IL-5, IL-6, IL-7, IL-8, IL-10, IL-12p40, IL-12p70, IL-13, IL-15, IL-17A, IL-17E/IL-25, IL-17F, IL-18, IL-22, IP-10, MCP-1, M-CSF, MIG, MIP-1α, MIP-1β, PDGF-AA, PDGF-AB/BB, TNF-α, TNF-β, and VEGF-A **[7]**.

### Statistical analysis

Categorical variables were evaluated using Fisher’s exact test. For clinical characteristics and cytokine data that deviated from normal distribution, the non-parametric Mann–Whitney U test was employed. To account for multiple cytokine comparisons, p-values were adjusted using the Benjamini–Hochberg procedure, with a false discovery rate (FDR)-adjusted q-value of ≤ 0.05 considered statistically significant.

### Data preprocessing

To ensure comparability across variables with differing scales, the data analysis process began with preprocessing to standardize all features. Specifically, z-score normalization **[13]** was used to transform each feature to have a mean of 0 and a standard deviation of 1, ensuring that all variables contributed equally to the model. This process is represented as follows:

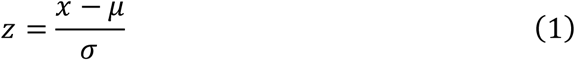

where *z* is the standardized value; *x* is the original value; *μ* is the mean of the feature; and *σ* is the standard deviation of the feature.

### Hierarchical clustering

Hierarchical clustering was used to group similar data elements into clusters and visualize them. The clustering results were represented using a dendrogram, a two-dimensional tree-like diagram depicting nested clusters **[14]**. Ward’s linkage method was applied **[15]**, which minimizes within-cluster variance, and Euclidean distance was used as the dissimilarity metric between variables and patients. The normalized data from preprocessing served as input for the clustering process.

In this study, hierarchical cluster analysis and heatmap generation were performed to investigate relationships among clinical parameters, cytokine profiles, and patient survival outcomes. In the heatmap, rows represented measured parameters (clinical characteristics and cytokines), while columns corresponded to individual patients, color-coded by outcome (green for survival and orange for mortality). Hierarchical clustering was applied to organize the rows based on expression pattern similarity across patients. The dendrogram on the left side of the heatmap depicted the clustering hierarchy, offering insights into parameters with shared expression patterns and potential biological relevance.

### Machine learning method

This section outlines the machine learning methods used in this study. The overall analytical workflow is depicted in **Figure 1**.

**Figure 1:**
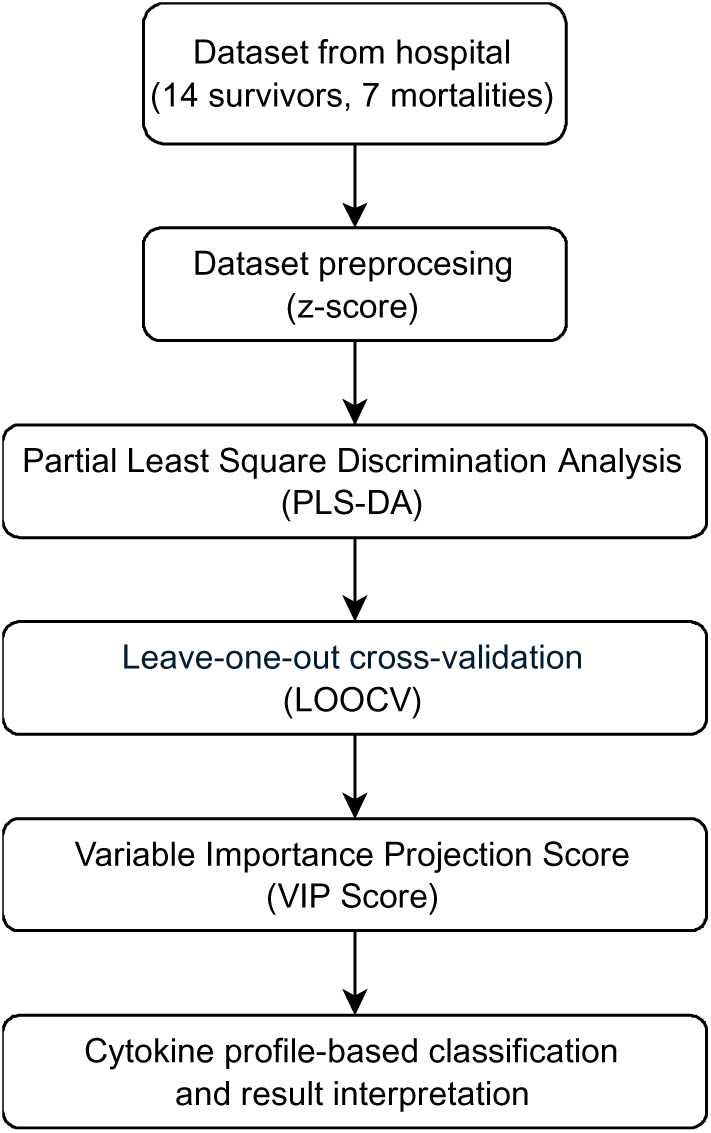
Workflow summary of the machine learning classification models.

### PLS-DA

PLS-DA was used to reduce the dataset’s high dimensionality and classify patients into survival and mortality groups. This technique is well-suited for datasets with correlated variables, as it projects the data onto latent variables that capture variance and outcome-relevant patterns. In doing so, PLS-DA reduces dimensionality while identifying features most critical for differentiating patient outcomes. This makes it particularly valuable for pinpointing key cytokines and clinical parameters linked to prognosis.

Given the sensitivity of PLS-DA to outliers, a robust version of the model was employed **[16]**. Outliers frequent in biomedical datasets due to patient heterogeneity can distort standard PLS-DA projections. To address this, a weighting mechanism based on residuals and leverage was applied, minimizing the impact of extreme values. This improved the accuracy and reliability of group classification and feature selection, especially for cytokine data, which are prone to biological and technical variability.

### Model evaluation

To validate the classification model, leave-one-out cross-validation (LOOCV) **[17]** was used to evaluate the performance of each feature subset. LOOCV provides a rigorous assessment by iteratively training the model on all samples except one and using the excluded sample to evaluate predictive performance. This method is well-suited for small datasets, as it maximizes data utilization while offering an unbiased performance estimate. LOOCV also helps mitigate overfitting and supports robust identification of cytokines associated with survival and mortality outcomes.

To evaluate the final model’s overall performance, receiver operating characteristic (ROC) curve analysis was applied. The ROC curve visually represents diagnostic ability by plotting the true positive rate against the false positive rate across classification thresholds. The area under the ROC curve (AUC-ROC) was calculated as a quantitative index of discriminative power. The AUC-ROC score ranges from 0.5 (random classification) to 1.0 (perfect classification), with higher values reflecting superior performance. This metric enables evaluation of the model’s ability to differentiate between survival and mortality across various decision thresholds, offering a comprehensive perspective on predictive accuracy.

In addition to the AUC-ROC, the model was further evaluated using traditional machine learning performance metrics, including accuracy, precision, recall, specificity, and F_1_ score. These metrics were defined based on the number of true positives (TP), false positives (FP), true negatives (TN), and false negatives (FN), as follows:

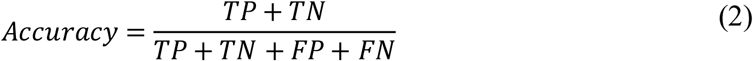

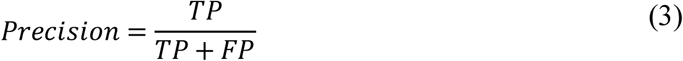

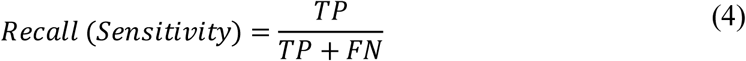

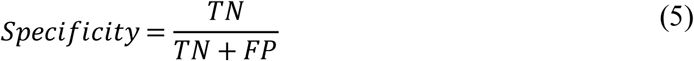

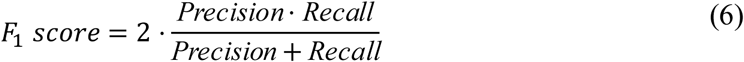

### Feature importance analysis

The VIP score was used to identify the most relevant cytokines for distinguishing between survival and mortality outcomes. The VIP score quantifies the contribution of each variable in the PLS-DA model also applied in this study. These scores are derived from the weighted sum of squared correlations between cytokine levels and the PLS-DA components:

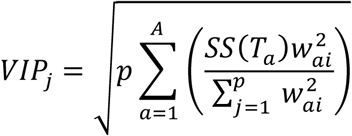

where *VIP*_*j*_ is the VIP score for feature *j*; *p* is the total number of features; *A* is the number of PLS components (latent variables); *w*_*ai*_ is the weight of feature *j* in component a, *SS*(*T*_*a*_) represents the variance in the response variable explained by component *a*; *and* 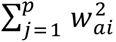 is the sum of the squared weights of all features for component *a*.

Features with higher VIP scores were considered more important for distinguishing between the survival and mortality groups. Typically, features with VIP scores greater than 1 were regarded as significant contributors to the model **[8]**. This approach enabled the identification of key cytokines that play a critical role in predicting survival outcomes, providing insight into the inflammatory processes underlying pediatric FM. The directionality of associations (positive or negative) was determined by the sign of the PLS regression coefficients. In this binary classification task, coefficients were extracted from the first latent component (PLS1), which best reflected the separation between survival and mortality groups. This made the interpretation of each cytokine’s role more straightforward and reliable. The combined use of VIP scores and PLS coefficients highlighted cytokines and clinical markers critical for predicting survival, offering mechanistic insight into dysregulated inflammatory pathways in pediatric FM.

## Results

### Statistical comparison of clinical characteristics and cytokine profile in survival and mortality groups

Baseline demographic characteristics and laboratory test results for the survival (n=14) and mortality (n=7) groups are summarized in **Table 1**. No statistically significant differences were observed in demographic parameters between the groups. Notably, sex, age, troponin T, and LVEF showed no significant differences despite clinical relevance: male sex (survival: 28.6% vs mortality: 42.9%, p=0.638), age (survival: 113 months [2–167] vs mortality: 150 months [14–180], p=0.636), troponin T (survival: 1.96 ng/mL [0.33–9.35] vs mortality: 2.72 ng/mL [0–24.7], p=0.971), and LVEF (survival: 30.7% [9–72] vs mortality: 10.7% [8–40], p=0.117). Arterial blood pH was the only clinical parameter that differed significantly between groups. The mortality group exhibited significantly lower pH than that of the survival group (mortality: 7.02 [6.56–7.35] vs survival: 7.33 [7.06–7.6]; p < 0.05) (**Table 1**)

**Table 1:**
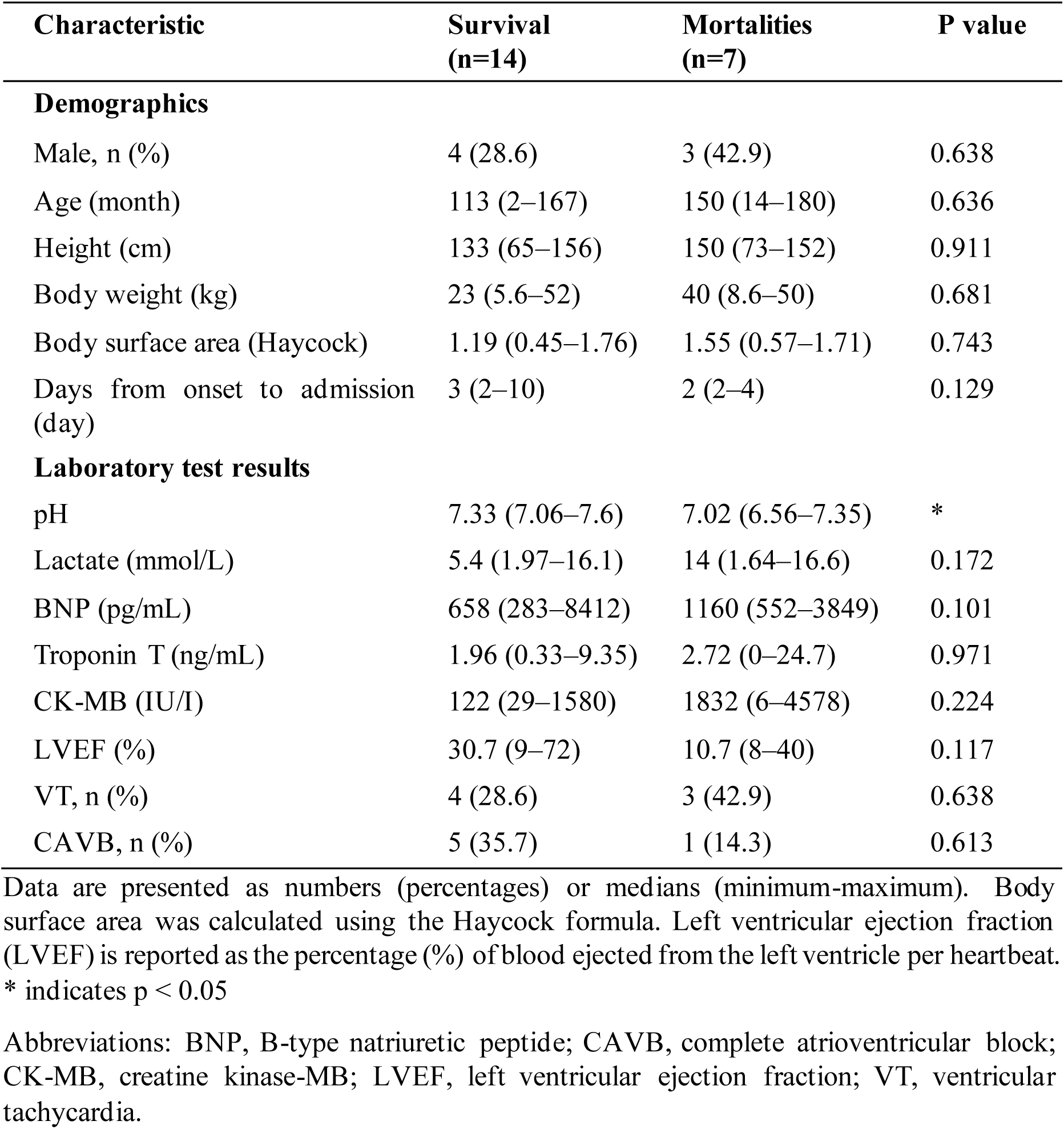
Demographic and clinical characteristics of patients with pediatric myocarditis.

| **Characteristic** | **Survival**<br>**(n=14)** | **Mortalities**<br>**(n=7)** | **P value** |
| --- | --- | --- | --- |
| **Demographics** |  |  |  |
| Male, n (%) | 4 (28.6) | 3 (42.9) | 0.638 |
| Age (month) | 113 (2–167) | 150 (14–180) | 0.636 |
| Height (cm) | 133 (65–156) | 150 (73–152) | 0.911 |
| Body weight (kg) | 23 (5.6–52) | 40 (8.6–50) | 0.681 |
| Body surface area (Haycock) | 1.19 (0.45–1.76) | 1.55 (0.57–1.71) | 0.743 |
| Days from onset to admission (day) | 3 (2–10) | 2 (2–4) | 0.129 |
| **Laboratory test results** |  |  |  |
| pH | 7.33 (7.06–7.6) | 7.02 (6.56–7.35) | * |
| Lactate (mmol/L) | 5.4 (1.97–16.1) | 14 (1.64–16.6) | 0.172 |
| BNP (pg/mL) | 658 (283–8412) | 1160 (552–3849) | 0.101 |
| Troponin T (ng/mL) | 1.96 (0.33–9.35) | 2.72 (0–24.7) | 0.971 |
| CK-MB (IU/I) | 122 (29–1580) | 1832 (6–4578) | 0.224 |
| LVEF (%) | 30.7 (9–72) | 10.7 (8–40) | 0.117 |
| VT, n (%) | 4 (28.6) | 3 (42.9) | 0.638 |
| CAVB, n (%) | 5 (35.7) | 1 (14.3) | 0.613 |

Data are presented as numbers (percentages) or medians (minimum-maximum). Body surface area was calculated using the Haycock formula. Left ventricular ejection fraction (LVEF) is reported as the percentage (%) of blood ejected from the left ventricle per heartbeat.
\* indicates $p < 0.05$
Abbreviations: BNP, B-type natriuretic peptide; CAVB, complete atrioventricular block; CK-MB, creatine kinase-MB; LVEF, left ventricular ejection fraction; VT, ventricular tachycardia.

Regarding the cytokine profile, seven cytokines exhibited statistically significant differences between the survival and mortality groups (q < 0.05). These included TNF-α (survival: 31.5 pg/mL [3.05–75.7] vs mortality: 91.5 pg/mL [37.0–168]), M-CSF (survival: 170 pg/mL [0–1,246] vs mortality: 1,787 pg/mL [254–3,625]), MIP-1α (survival: 23.8 pg/mL [9.82–53.5] vs mortality: 52.1 pg/mL [31.2–64.8]), IL-8 (survival: 26.8 pg/mL [8.69–197] vs mortality: 372 pg/mL [50.6–1,363]), IL-6 (survival: 22.5 pg/mL [0.24–579] vs mortality: 1,862 pg/mL [32.7–10,404]), IL-15 (survival: 10.8 pg/mL [3.29–69.9] vs mortality: 31.2 pg/mL [18.2–85.0]), and IP-10 (survival: 683 pg/mL [117–11,374] vs mortality: 21,636 pg/mL [1,269–9,536,904])(**Table 2**).

**Table 2:**
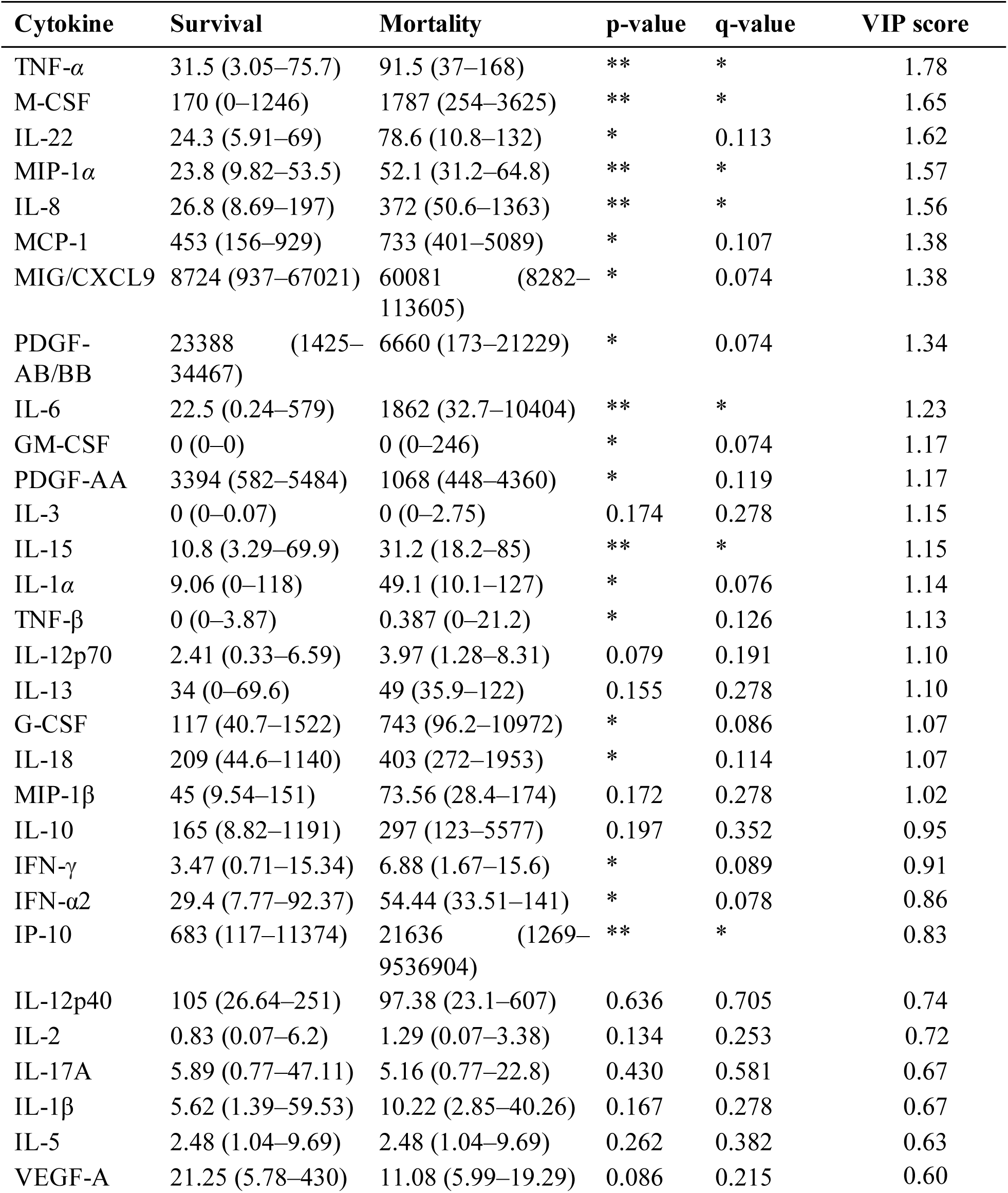
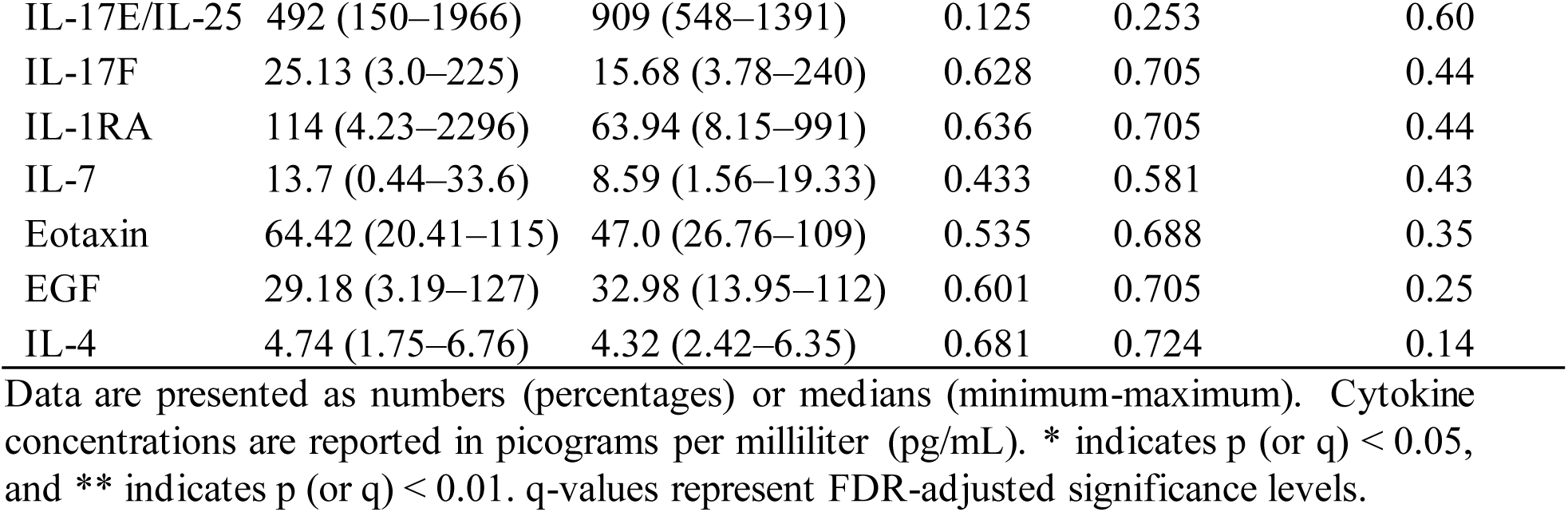
Comparison of cytokine levels in survival and mortality groups.

| **Cytokine** | **Survival** | **Mortality** | **p-value** | **q-value** | **VIP score** |
| --- | --- | --- | --- | --- | --- |
| TNF-α | 31.5 (3.05–75.7) | 91.5 (37–168) | ** | * | 1.78 |
| M-CSF | 170 (0–1246) | 1787 (254–3625) | ** | * | 1.65 |
| IL-22 | 24.3 (5.91–69) | 78.6 (10.8–132) | * | 0.113 | 1.62 |
| MIP-1α | 23.8 (9.82–53.5) | 52.1 (31.2–64.8) | ** | * | 1.57 |
| IL-8 | 26.8 (8.69–197) | 372 (50.6–1363) | ** | * | 1.56 |
| MCP-1 | 453 (156–929) | 733 (401–5089) | * | 0.107 | 1.38 |
| MIG/CXCL9 | 8724 (937–67021) | 60081 (8282–113605) | * | 0.074 | 1.38 |
| PDGF-AB/BB | 23388 (1425–34467) | 6660 (173–21229) | * | 0.074 | 1.34 |
| IL-6 | 22.5 (0.24–579) | 1862 (32.7–10404) | ** | * | 1.23 |
| GM-CSF | 0 (0–0) | 0 (0–246) | * | 0.074 | 1.17 |
| PDGF-AA | 3394 (582–5484) | 1068 (448–4360) | * | 0.119 | 1.17 |
| IL-3 | 0 (0–0.07) | 0 (0–2.75) | 0.174 | 0.278 | 1.15 |
| IL-15 | 10.8 (3.29–69.9) | 31.2 (18.2–85) | ** | * | 1.15 |
| IL-1α | 9.06 (0–118) | 49.1 (10.1–127) | * | 0.076 | 1.14 |
| TNF-β | 0 (0–3.87) | 0.387 (0–21.2) | * | 0.126 | 1.13 |
| IL-12p70 | 2.41 (0.33–6.59) | 3.97 (1.28–8.31) | 0.079 | 0.191 | 1.10 |
| IL-13 | 34 (0–69.6) | 49 (35.9–122) | 0.155 | 0.278 | 1.10 |
| G-CSF | 117 (40.7–1522) | 743 (96.2–10972) | * | 0.086 | 1.07 |
| IL-18 | 209 (44.6–1140) | 403 (272–1953) | * | 0.114 | 1.07 |
| MIP-1β | 45 (9.54–151) | 73.56 (28.4–174) | 0.172 | 0.278 | 1.02 |
| IL-10 | 165 (8.82–1191) | 297 (123–5577) | 0.197 | 0.352 | 0.95 |
| IFN-γ | 3.47 (0.71–15.34) | 6.88 (1.67–15.6) | * | 0.089 | 0.91 |
| IFN-α2 | 29.4 (7.77–92.37) | 54.44 (33.51–141) | * | 0.078 | 0.86 |
| IP-10 | 683 (117–11374) | 21636 (1269–9536904) | ** | * | 0.83 |
| IL-12p40 | 105 (26.64–251) | 97.38 (23.1–607) | 0.636 | 0.705 | 0.74 |
| IL-2 | 0.83 (0.07–6.2) | 1.29 (0.07–3.38) | 0.134 | 0.253 | 0.72 |
| IL-17A | 5.89 (0.77–47.11) | 5.16 (0.77–22.8) | 0.430 | 0.581 | 0.67 |
| IL-1β | 5.62 (1.39–59.53) | 10.22 (2.85–40.26) | 0.167 | 0.278 | 0.67 |
| IL-5 | 2.48 (1.04–9.69) | 2.48 (1.04–9.69) | 0.262 | 0.382 | 0.63 |
| VEGF-A | 21.25 (5.78–430) | 11.08 (5.99–19.29) | 0.086 | 0.215 | 0.60 |
| IL-17E/IL-25 | 492 (150–1966) | 909 (548–1391) | 0.125 | 0.253 | 0.60 |
| IL-17F | 25.13 (3.0–225) | 15.68 (3.78–240) | 0.628 | 0.705 | 0.44 |
| IL-1RA | 114 (4.23–2296) | 63.94 (8.15–991) | 0.636 | 0.705 | 0.44 |
| IL-7 | 13.7 (0.44–33.6) | 8.59 (1.56–19.33) | 0.433 | 0.581 | 0.43 |
| Eotaxin | 64.42 (20.41–115) | 47.0 (26.76–109) | 0.535 | 0.688 | 0.35 |
| EGF | 29.18 (3.19–127) | 32.98 (13.95–112) | 0.601 | 0.705 | 0.25 |
| IL-4 | 4.74 (1.75–6.76) | 4.32 (2.42–6.35) | 0.681 | 0.724 | 0.14 |

Data are presented as numbers (percentages) or medians (minimum-maximum). Cytokine concentrations are reported in picograms per milliliter (pg/mL). \* indicates $p$ (or $q$ ) < 0.05, and \*\* indicates $p$ (or $q$ ) < 0.01. $q$ -values represent FDR-adjusted significance levels.

### Hierarchical clustering of clinical parameters and cytokines

Further analysis using hierarchical clustering of clinical parameters and cytokines revealed distinct expression patterns between the survival and mortality groups (**Figure 2**). The accompanying dendrograms provided additional insight into relationships among cytokines and patient profiles. While most demographic and clinical parameters showed no statistically significant group differences, clustering of cytokine profiles suggested that certain patterns, particularly among variables clustered below Male, may reflect group-specific trends. Although these clusters did not clearly segregate the survival and mortality groups, they indicate latent multivariate differences that merit further investigation. To identify more precisely key predictive features, a machine learning approach (PLS-DA) was applied for quantitative validation of observed group differences.

**Figure 2:**
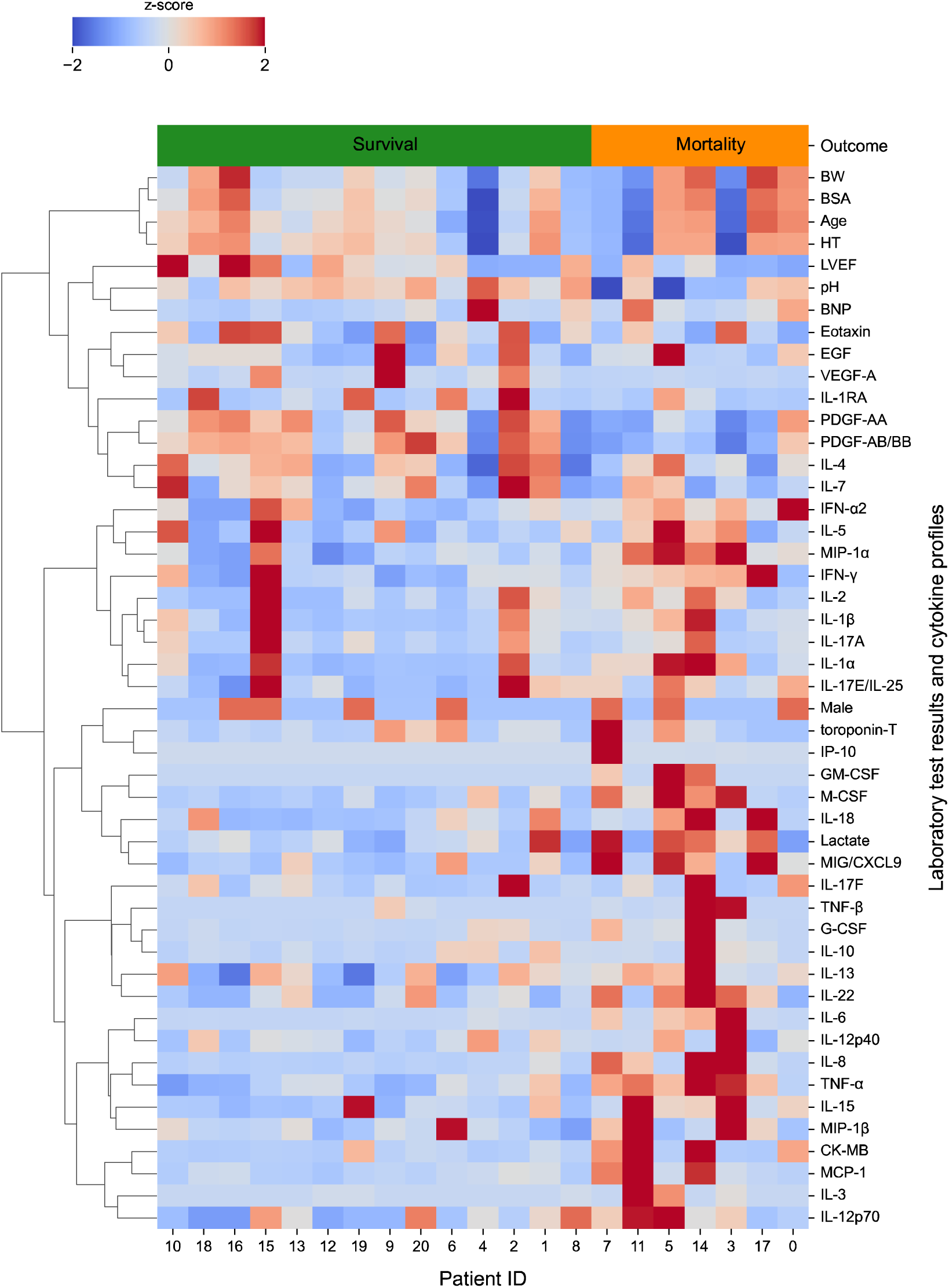
Hierarchical clustering heatmap. The top dendrogram illustrates the clustering of patients, while the side dendrogram displays the clustering of clinical characteristics and cytokines. The right side indicates the two groups (survival and mortality) analyzed. In the heatmap, each column represents an individual patient (color-coded by outcome: green for survival, orange for mortality), and each row represents a distinct clinical characteristic or cytokine, organized by expression similarity. Heatmap colors indicate cytokine expression intensities.

### PLS-DA in survival and mortality groups

Encouraged by additional statistical analysis, the PLS-DA model addressed multicollinearity among 51 features; 14 clinical variables (six demographic parameters and eight laboratory test results) and 37 cytokines. As shown in the score plot (**Figure 3a**), clear separation was achieved between the survival (n=7) and mortality (n=14) groups, with only one mortality case overlapping into the survival region, and most data points falling within their respective 95% confidence ellipses. The model was validated using LOOCV, achieving an overall accuracy of 85.7% (**Figure 3b**), with a sensitivity of 92.9% (13 of 14 non-survivors correctly identified) and specificity of 71.4% (five of seven survivors accurately classified). Misclassifications included two non-survivors predicted as survivors and one survivor classified as a non-survivor. ROC analysis yielded an AUC of 0.90 (**Figure 3c**), with a steep initial curve trajectory.

**Figure 3:**
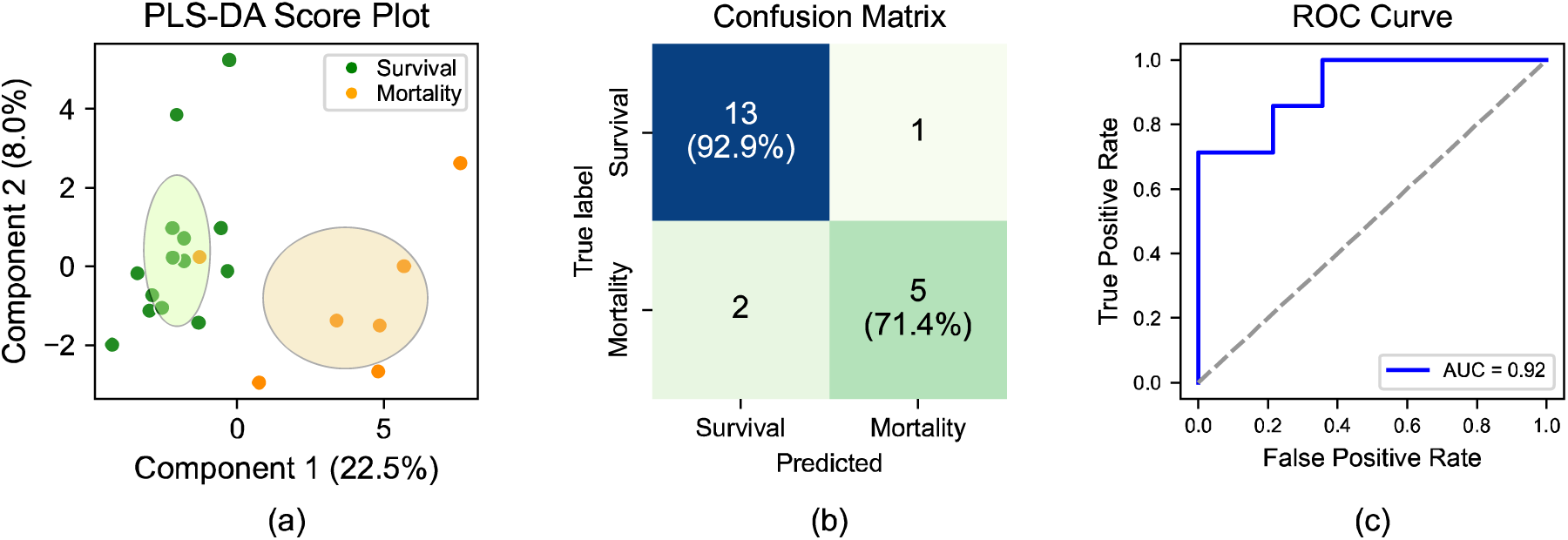
Partial least squares discriminant analysis (PLS-DA) results. (a) PLS-DA score plot illustrating the separation between survival (green) and mortality (orange) groups. Each dot represents an individual patient. For each group, an ellipse is overlaid to depict score dispersion. These ellipses are centered at the group means, with their widths and heights set to twice the standard deviation of the scores along the respective axes. (b) Confusion matrix presenting counts and percentages for predictions. (c) ROC curve with AUC, indicating the model’s classification performance.

The PLS-DA model identified 23 prognostic features with VIP scores > 1.0, indicating their significant contribution to mortality prediction in pediatric FM (**Figure 4**). These included three clinical laboratory variables: pH (p<0.05, VIP=1.45), lactate (p=0.172, VIP=1.25), and CK-MB (p=0.224, VIP=1.21), as well as 20 of 37 cytokines. All selected cytokines demonstrated distinct expression patterns between survivors and non-survivors, as visualized in the boxplots in **Figure 5**.

**Figure 4:**
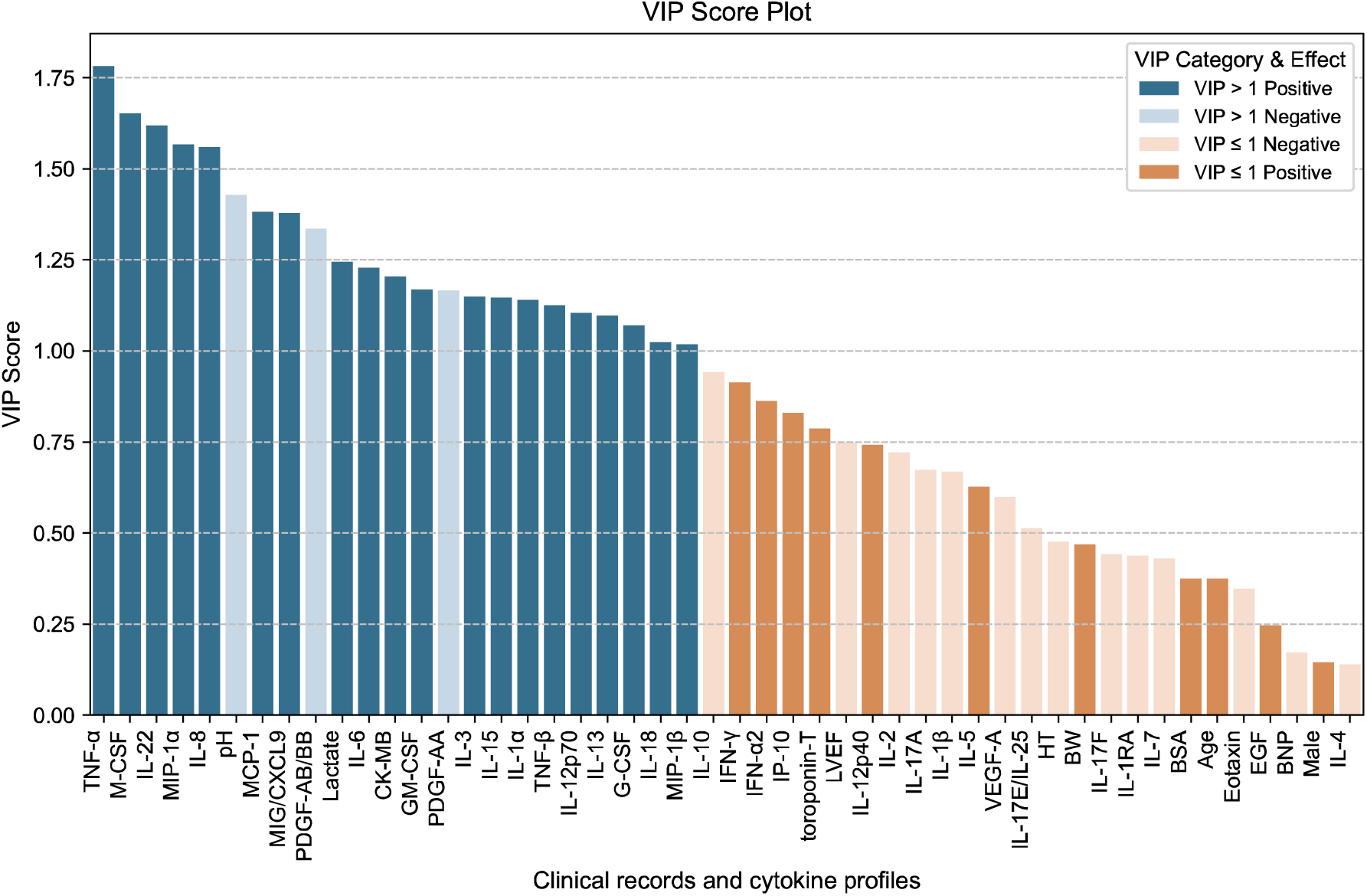
Summary of variable importance in projection (VIP) scores for features analyzed in the study. Features with VIP scores greater than or equal to 1 are displayed in the left columns, indicating greater importance in classification. Features with VIP scores less than 1 are shown in the right columns. VIP scores were derived from the partial least squares discriminant analysis (PLS-DA) model. Directionality (positive/negative) indicates each feature’s association with mortality: a positive effect reflects a higher level of the feature associated with increased mortality risk, whereas a negative effect indicates a lower level associated with mortality.

**Figure 5:**
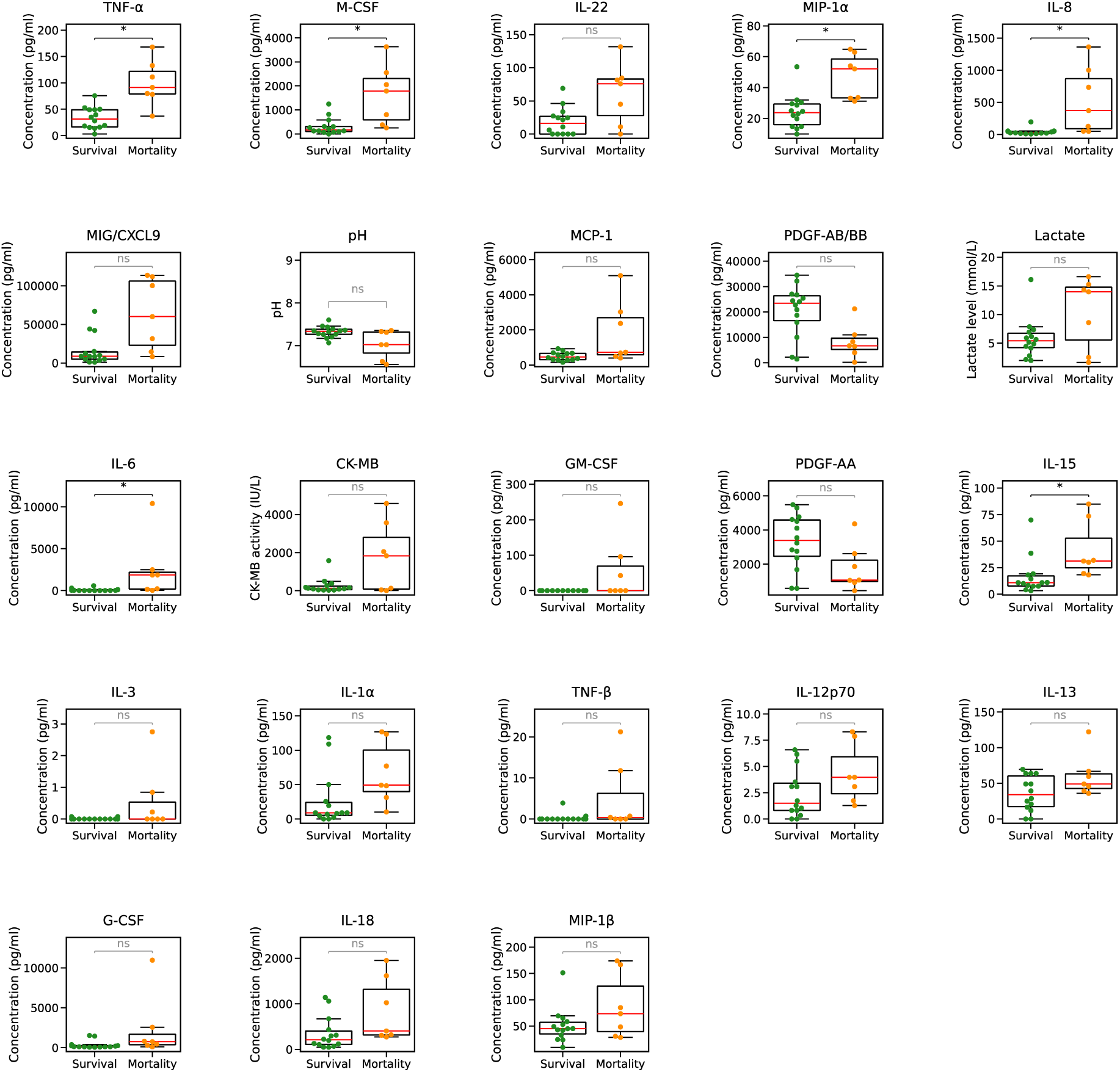
Box-and-whisker plots combined with scatter plots, comparing features between survival and mortality groups. Only features with VIP scores greater than 1 are shown. * indicates p<0.05, ** indicates p<0.01, and ns indicates non-significant differences.

Among the clinical variables, lactate and CK-MB showed positive VIP score contributions, indicating elevated levels in non-survivors, while pH demonstrated a negative VIP score contribution, corresponding to significantly reduced levels in the mortality group (**Figure 4**). Cytokine profiling revealed six dual-significance biomarkers that met both PLS-DA importance criteria (VIP > 1.0) and FDR-adjusted statistical significance (q < 0.05): TNF-α (VIP=1.78), M-CSF (VIP=1.65), MIP-1α (VIP=1.57), IL-8 (VIP=1.56), IL-6 (VIP=1.23), and IL-15 (VIP = 1.15). An additional 14 cytokines demonstrated high importance in the PLS-DA model (VIP > 1.0) but did not reach univariate statistical significance after FDR correction (**Table 2**), including IL-22, MCP-1, MIG/CXCL9, GM-CSF, IL-3, IL-1α, TNF-β, IL-12p70, IL-13, G-CSF, IL-18, and MIP-1β. Notably, PDGF-AA and PDGF-AB/BB, the only cytokines inversely associated with mortality, showed reduced levels in non-survivors (**Figure 4**) and negative VIP score contributions (**Figure 5**), while all other cytokines exhibited positive associations with mortality. The complete VIP score distribution and associated statistical results from the PLS-DA model are presented in **Figure 4**.

## Discussions

In this study, a machine learning model was applied to analyze cytokine profiles and clinical features associated with mortality in FM. The results indicated that several cytokines, including TNF-α, along with clinical parameters such as pH, lactate, and CK-MB, were significantly associated with mortality.

Our analysis of cytokine profiles in FM identified elevated IL-6, IL-8, and IL-15, and reduced PDGF-AA as cytokines linked to mortality. These cytokines, previously examined in our study stratifying AM and FM **[7]**, also appear to be related to mortality. Our findings suggest that cytokines contributing to disease severity in AM may overlap with those associated with poor outcomes in FM.

In our latest machine learning-based analysis, serum cytokines measured at admission, particularly TNF-α, emerged as key factors associated with mortality in FM. Notably, TNF-α was not the primary focus of our previous cytokine studies aimed at stratifying AM severity **[7]**. Although prior studies have linked TNF-α to FM pathophysiology, paradoxically, TNF-α suppression has been reported to delay healing and worsen disease outcomes **[18]**. However, recent animal model studies on FM have highlighted the critical role of TNF-α in the early inflammatory response, showing that the timing of TNF-α inhibition is pivotal. Specifically, targeted suppression of TNF-α at appropriate disease stages has demonstrated improved outcomes **[19]**. Currently, treatment strategies for FM do not include infliximab, an anti-TNF-α agent **[1]**. Given the severe circulatory failure requiring MCS observed in early FM cases, anti-TNF-α therapy may help resolve intense myocardial inflammation earlier, potentially improving outcomes. However, further clinical investigations in humans are needed to validate this hypothesis and explore the therapeutic role of targeted TNF-α inhibition in FM.

VIP score analysis revealed that troponin T was not significantly associated with mortality, whereas CK-MB demonstrated a significant correlation with fatal outcomes. Previous studies have similarly reported that troponin is not a risk factor for cardiac arrest or the need for MCS in pediatric myocarditis **[4]**. FM is characterized by a rapid transition from initial flu-like symptoms to circulatory collapse, often resulting in fatal outcomes without timely therapeutic intervention **[20]**. Notably, the time from symptom onset to hospitalization is approximately 3 days **[21]**, a finding consistent with our current analysis. This supports the notion that FM follows a predictable progression. Regarding the kinetics of myocardial injury markers, high-sensitivity troponin rises earlier, followed by CK-MB. Troponin levels typically normalize within 7–10 days. Due to its shorter half-life, CK-MB reaches peak levels concurrent with the onset of acute circulatory failure **[22]**. These findings suggest that CK-MB may be a sensitive biomarker for predicting early mortality in FM. Moreover, a 2024 study using myocardial cells from FM patients demonstrated that CK-MB showed a stronger correlation with the severity of acute myocardial injury than did troponin **[23]**. This provides strong supporting evidence for our findings, reinforcing the prognostic utility of CK-MB in FM.

LVEF, an echocardiographic parameter reflecting systolic function, was not associated with mortality in the VIP score analysis. In contrast, the study by Zhao et al. identified LVEF as a prognostic predictor in FM **[3]**. However, direct comparisons between studies are limited by differences in diagnostic criteria and the accuracy of FM diagnosis. In a study on cardiogenic shock in adults, no statistically significant difference in LVEF was observed between individuals with and without cardiac arrest **[24]**. Similarly, LVEF may not be a reliable prognostic marker in FM, as previously reported in pediatric myocarditis, where it was not associated with cardiac arrest or the need for MCS **[4]**. LVEF is known to be highly variable due to its sensitivity to preload and afterload conditions, making accurate assessment difficult in the context of cardiogenic shock and high-dose catecholamine therapy. In our study, in addition to cases with irreversible myocardial damage and severe systolic dysfunction requiring ECMO support, fatal arrhythmias such as CAVB also contributed to the ECMO indication. Therefore, factors beyond systolic function may have contributed to circulatory failure, potentially influencing our results. Further investigation is warranted to clarify the prognostic significance of LVEF in FM

In this study, although lactate levels did not show a statistically significant difference between the survival and mortality groups, they were identified as a prognostic feature with a high VIP score in the machine learning model. Clinically, lactate, like pH, is widely used as a key indicator of hemodynamic status **[3]**. These findings suggest that parameters emphasized by clinicians through experience or intuition may not reach significance in univariate analyses but can be effectively captured and weighted in machine learning models. Integrating clinical insights with objective machine learning analysis allows for the development of predictive models that are more relevant in real-world settings. Ongoing collaboration between clinical experts and data scientists will be essential in future research to identify clinically meaningful predictors and enhance the validity and utility of prognostic models.

A similar pattern was observed for IP-10, which demonstrated a statistically significant difference between groups (q < 0.05); however, it was not identified as an important variable by the machine learning model, as indicated by its VIP score of 0.83. This discrepancy may reflect a limitation of the PLS-DA model, which often selects only one variable among those with highly correlated data patterns. Notably, the expression pattern of IP-10 was strongly correlated with that of TNF-α (correlation coefficient=0.89), suggesting potential redundancy in the model’s feature selection. IP-10 has been previously implicated in the pathogenesis of myocarditis **[25–27]**, and murine model studies have shown that suppression of endogenous IP-10 leads to reductions in serum CK-MB levels and improved survival outcomes **[28]**. These findings underscore the potential of IP-10 as a therapeutic target for future anti-cytokine interventions. Moving forward, the development of more accurate prognostic models will require refinement of machine learning approaches and expanded collection of serum cytokine profiles, particularly in individuals with FM.

In the analysis of cytokine profiles and clinical data, IL-22, MCP-1, MIG/CXCL9, PDGF-AB/BB, GM-CSF, PDGF-AA, IL-3, IL-1α, TNF-β, IL-12p70, IL-13, G-CSF, IL-18, MIP-1β, CK-MB, and lactate did not exhibit statistically significant differences in conventional univariate analyses. However, machine learning identified these factors as being associated with mortality in FM. Notably, many of these cytokines have previously been implicated in the pathogenesis of myocarditis **[29–36]**. While traditional statistical tests assess the presence or absence of differences in individual variables, machine learning evaluates the collective discriminatory capacity of multiple features. This suggests that these factors may not independently distinguish FM mortality but rather reflect complex, interdependent biological interactions characteristic of myocarditis. Machine learning-based analyses, therefore, hold promise for uncovering novel biomarkers that might be overlooked by conventional methods. Future studies employing these advanced techniques could yield valuable insights into the pathophysiology of FM and inform the development of targeted clinical interventions to improve outcomes for affected individuals.

## Limitations

First, this study employed a retrospective design, which introduces inherent constraints in data collection and interpretation. Retrospective studies are prone to inconsistencies in data completeness, as clinical records may not have been collected uniformly. Moreover, therapeutic interventions and disease trajectories cannot be evaluated in real time, necessitating cautious interpretation of causal relationships compared to prospective approaches. Second, an imbalance in the dataset posed a key limitation. The significantly higher number of survivors than non-survivors may have reduced recall and sensitivity, particularly in mortality prediction. The small number of non-survivor cases may have limited the model’s capacity to learn discriminative patterns, potentially resulting in an underestimation of mortality risk. However, pediatric FM is an extremely rare condition, and data collection is inherently challenging due to its low prevalence and the need for specialized clinical settings. Third, although the exact timing of FM onset remains uncertain in many cases, variations in disease progression are likely minimal, as the condition generally follows a consistent clinical course. Therefore, discrepancies in the timing of cytokine and clinical marker measurement are unlikely to influence study outcomes meaningfully. Nevertheless, the precise timing of myocardial injury marker elevations (e.g., CK-MB and troponin) in early-stage FM remains unclear and should be investigated in future studies.

## Conclusions

This study highlights the potential of machine learning models to predict outcomes in pediatric FM by integrating clinical features and cytokine profiling. The observed associations between cytokines, particularly TNF-α, and prognosis support the need for further investigation into anti-cytokine therapeutic strategies for managing pediatric FM.

## Non-standard Abbreviations and Acronyms

AM: Acute Myocarditis
AUC-ROC: Area Under the Receiver Operating Characteristic Curve
BNP: B-type Natriuretic Peptide
CAVB: Complete Atrioventricular Block
CK-MB: Creatine Kinase MB
ECMO: Extracorporeal Membrane Oxygenation
FDR: False Discovery Rate
FM: Fulminant Myocarditis
FN: False Negatives
FP: False Positives
LOOCV: Leave-One-Out Cross-Validation
LVEF: Left Ventricular Ejection Fraction
MCS: Mechanical Circulatory Support
PLS-DA: Partial Least Square Discriminant Analysis
ROC: Receiver Operating Characteristic Curve
TNF-α: Tumor necrosis factor-alpha
TN: True Negatives
TP: True Positives
VIP: Variable Importance in Projection
VT: Ventricular Tachycardia

## Acknowledgments

We extend our sincere appreciation to Naoko Takahashi at the Aichi Children’s Health and Medical Center for her invaluable assistance with specimen collection; to Akiko Yoshikawa at Fujita Health University School of Medicine for her technical support during experimental procedures; to Toshiya Kokaji at the Nara Institute of Science and Technology for his insightful and technical guidance on statistical analyses; and to Aamir Jeewa at the Hospital for Sick Children and Editage (www.editage.com) for their content review and contributions to the English-language refinement.

## ETHICS APPROVAL

This study was approved by the institutional review boards of Aichi Children’s Health and Medical Center and Fujita Health University (No. 2019027 and HM21-575). All procedures were conducted in accordance with the ethical guidelines of Institution and the Declaration of Helsinki. Informed consent was obtained from all participants prior to their involvement in the study.

## DATA AVAILABILITY

De-identified data analyzed in this study are presented in the main text and tables. Additional data are available from the corresponding author upon reasonable request.

## AUTHOR CONTRIBUTIONS

SJ, TS, YN, KK, YS, KTS, AF and KY conceptualized and designed the study, conducted data analyses, drafted the initial manuscript, and contributed to critical revisions. TS, YN, and HU collected clinical data. SJ, TS, KK, YS, and KTS contributed to bioinformatics and clinical data interpretation. TS and JY secured funding for the project. All authors reviewed and approved the final manuscript and agreed to be accountable for all aspects of the work.

